# Modeling lesion transition dynamics to clinically characterize mpox patients in the Democratic Republic of the Congo

**DOI:** 10.1101/2024.01.28.24301907

**Authors:** Takara Nishiyama, Fuminari Miura, Yong Dam Jeong, Naotoshi Nakamura, Hyeongki Park, Masahiro Ishikane, Shotaro Yamamoto, Noriko Iwamoto, Michiyo Suzuki, Ayana Sakurai, Kazuyuki Aihara, Koichi Watashi, William S Hart, Robin N Thompson, Yasuhiro Yasutomi, Norio Ohmagari, Placide Mbala Kingebeni, John W. Huggins, Shingo Iwami, Phillip R. Pittman

## Abstract

Coinciding with the global outbreak of clade IIb mpox virus (MPXV), the Democratic Republic of the Congo (DRC) recently experienced a rapid surge in mpox cases with clade I MPXV. Clade I MPXV is known to be more fatal, but its clinical characteristics and prognosis differ between patients. Here, we used mathematical modelling to quantify disease progression in a large cohort of mpox patients in the DRC from 2007-2011, particularly focusing on lesion transition dynamics. We further analyzed individuals’ clinical data to find predictive biomarkers of severity of symptoms. Our analysis shows that mpox patients can be stratified into three groups according to symptom severity, and that viral load at symptom onset may serve as a predictor to distinguish groups with the most severe or mild symptoms after progression. Understanding the severity and duration of symptoms in different patients, as characterized by our approach, allows treatment strategies to be improved and individual-specific control measures (e.g isolation strategies based on disease progression) to be developed.

## Main Text

Mpox (formerly known as monkeypox) is a zoonotic disease caused by the monkeypox virus (MPXV), resulting in a rash [1-3]. MPXV was first discovered in humans in the Democratic Republic of the Congo (DRC) in 1970 and is classified into two clades (clade I and clade II). Both clades have been endemic in countries in Central and West Africa, spreading mainly by animal-to-human transmission [4-8]. A recent global mpox outbreak with clade IIb emerged through human-to-human transmission in Western countries, predominantly affecting men who have sex with men via sexually associated transmission, starting in May 2022 [9-12]. Initially rapid, the mpox outbreak was no longer considered a public health emergency of international concern (PHEIC) by May 2023 [13]. Nevertheless, following earlier concerns [14, 15], the number of mpox cases has significantly increased in the Western Pacific Region since April 2023, particularly in China. Moreover, the DRC has recently documented transmission via sexual contact of clade I MPXV [16], which has a case fatality ratio of about 10% [6, 7, 17, 18], significantly higher than that of clade IIb during 2022 (<1%) [19-22]. There has been a rapid increase in the number of suspected cases of clade I in the DRC (with more than 10,000 reported cases in 2023) [23], further raising concerns about the potential global spread of mpox, potentially with this more fatal clade, despite termination of the PHEIC due to clade IIb.

Understanding disease progression is key for both treatment and outbreak control. Common symptoms of mpox include a skin rash or mucosal lesions accompanied by fever, headache, muscle aches, back pain, low energy, and swollen lymph nodes [10, 22, 24-27]. The skin lesions associated with mpox typically persist for 2 to 3 weeks and progress through seven stages: macules, papules, vesicles, and then pustules, followed by umbilication, scabbing, and desquamation [28-30]. Current recommendations for patients are to refrain from direct skin contact from the symptom onset until lesions have healed and scabs fall off (i.e., a symptom-based isolation rule [31]), given that person-to-person transmission of mpox can occur through direct contact with infectious skin or other lesions [28-30]. Notably, the viral load in skin samples is significantly higher, by several orders of magnitude, and persists longer than the viral load in other body locations [32, 33]. The number of skin lesions that a patient experiences during MPXV infection varies over time and between individuals, ranging from a few to over 1000 [8, 25, 27]. Clinical features of the rash also differ depending on MPXV clade [26, 32, 34, 35]. Thus, individual-level dynamics in the number of lesions provide evidence for determining tailored treatment and effective interventions. However, mpox lesion dynamics have not previously been characterized quantitatively, and in particular, individual heterogeneity has yet to be fully examined.

In this study, we aimed to quantify the time course of disease progression of mpox and find predictors of individual-level severity of symptoms, using a large longitudinal dataset that contains lesion counts, viral loads of various specimens, and other biomarkers from patients infected with clade I MPXV in the DRC during 2007–2011 [27]. Our analysis stratified the study population into three groups by severity of lesion symptoms (e.g., mild or severe progressors), clinically characterized them, and identified clinical markers predicting severity. These findings have the potential to contribute to the development of effective treatment strategies and control measures, not only for clade I MPXV but also other MPXV clades and human orthopoxviruses [36].

## Results

### Description of cohort and study design

We used comprehensive clinical data collected from MPXV-infected patients at the remote L’Hopital General de Reference de Kole (Kole Hospital), in the rainforest of the Congo River basin of DRC, from March 2007 until August 2011. The data were collected as part of a study conducted jointly by the Institute National de Recherche Biomedical (INRB) and the US Army Medical Research Institute of Infectious Diseases (USAMRIID) [27].

Of 244 patients diagnosed with MPXV infection, 228 tested positive by MPXV-specific PCR testing. In addition, 13 patients were excluded because of incomplete observational data. In our analysis, we studied lesion transition dynamics in 149 participants and virus dynamics in 151 participants, excluding 66 and 64 individuals, respectively, owing to incomplete longitudinal data on lesion counts and viral load (see **Extended Data Fig 1**). Among the 228 patients who tested positive, 3 were fetal cases. The dataset comprised lesion counts at different stages on various areas of the patients’ bodies, as well as the MPXV genomic DNA load (i.e., viral load) in peripheral blood, pharyngeal swabs, and scabs from the same individuals. Additionally, we integrated clinical laboratory data from hematology analysis, urinalysis, and clinical chemistry within the same patient cohort; **Table 1** shows the summary statistics of these measured items for all individuals (total) and by the level of disease severity (G1, G2, and G3), which is characterized in the next section. Furthermore, the comprehensive clinical dataset for these patients included annotations for their sex, age, clinical symptoms, clinical signs, and exposure possibilities (details are outlined in [27]).

**Table 1.** Summary of clinical laboratory data of 149 mpox patients for lesion transition analysis.

|  | Total (N=149) | G1 (N=42) | G2 (N=31) | G3 (N=76) | Adjusted P value † |
| --- | --- | --- | --- | --- | --- |
| **Chemistry** |  |  |  |  |  |
| Glucose | 93.3 [20.9] | 87.3 [16.6]* | 95.9 [21.8] | 94.6 [22.6] | 1.000 |
| Blood urea nitrogen | 8.23 [5.22] | 6.18 [2.78] | 8.63 [4.97] | 10.1 [7.18] | 0.610 |
| Creatinine | 0.72 [0.50] | 0.84 [0.80] | 0.69 [0.30] | 0.62 [0.31] | 1.000 |
| Uric acid | 4.60 [1.80] | 4.44 [1.40] | 4.65 [1.95] | 4.71 [1.96] | 0.978 |
| Calcium | 8.82 [0.80] | 8.60 [1.29] | 8.87 [0.47] | 9.01 [0.52] | 1.000 |
| Albumin | 2.79 [0.55] | 2.89 [0.88] | 2.73 [0.38] | 2.83 [0.33] | 1.000 |
| Total protein | 7.72 [0.98] | 7.67 [1.33] | 7.73 [0.74] | 7.76 [0.99] | 1.000 |
| Alanine aminotransferase | 23.6 [10.7] | 21.4 [9.52] | 25.2 [11.9] | 22.7 [8.33] | 1.000 |
| Aspartate aminotransferase | 44.5 [22.4] | 39.5 [24.8] | 44.5 [20.1] | 51.3 [23.5] | 0.377 |
| Alkaline phosphatase | 118 [45.6] | 116 [52.1] | 117 [40.2] | 123 [49.9] | 0.942 |
| Total bilirubin | 0.62 [0.19] | 0.61 [0.20] | 0.60 [0.19] | 0.66 [0.20] | 1.000 |
| Gamma glutamyl transferase | 40.6 [36.9] | 32.4 [19.3] | 42.1 [40.0] | 48.0 [45.0] | 1.000 |
| Amylase | 79.3 [102] | 75.1 [40.1] | 82.0 [123] | 78.3 [100] | 1.000 |
| **Complete blood count** |  |  |  |  |  |
| Neutrophils | 38.8 [13.1] | 33.5 [12.8] | 39.1 [11.0] | 45.2 [15.7] | 0.080 |
| BANDS | 1.17 [2.98] | 0.65 [1.90] | 1.12 [2.46] | 1.97 [4.72] | 1.000 |
| Lymphocytes | 41.2 [15.7] | 46.7 [13.3] | 40.9 [15.9] | 34.6 [15.8] | 0.210 |
| Atypical lymphocytes | 0.78 [1.33] | 0.63 [1.25] | 0.91 [1.35] | 0.68 [1.40] | 1.000 |
| Metamyelocytes | 0.01 [0.08] | 0 [0] | 0.01 [0.12] | 0 [0] | 1.000 |
| Myelocytes | 0.27 [0.73] | 0.10 [0.38] | 0.33 [0.83] | 0.36 [0.80] | 1.000 |
| Monocytes | 5.55 [6.61] | 5.30 [6.84] | 6.17 [6.91] | 4.36 [5.51] | 1.000 |
| Eosinophils | 11.5 [6.48] | 12.8 [6.93] | 11.1 [5.89] | 10.6 [7.21] | 1.000 |
| Basophils | 0.29 [0.59] | 0.30 [0.46] | 0.23 [0.48] | 0.42 [0.89] | 1.000 |
| White blood cell | 10.7 [5.15] | 9.29 [4.26] | 10.4 [5.13] | 13.3 [5.44] | 0.017 |
| Red blood cell | 4.50 [0.71] | 4.45 [0.78] | 4.49 [0.67] | 4.61 [0.75] | 1.000 |
| Hemoglobin | 11.6 [2.02] | 11.4 [2.22] | 11.6 [1.83] | 11.8 [2.31] | 1.000 |
| Hematocrit | 34.7 [5.84] | 34.5 [6.33] | 34.6 [5.55] | 35.1 [6.02] | 1.000 |
| Mean corpuscular volume | 77.9 [7.72] | 79.1 [8.99] | 77.6 [7.23] | 77.1 [7.25] | 1.000 |
| Mean corpuscular hemoglobin | 25.9 [2.85] | 25.8 [3.24] | 25.9 [2.68] | 25.6 [2.88] | 1.000 |
| Mean corpuscular hemoglobin concentration | 33.2 [1.89] | 32.6 [1.64] | 33.5 [2.08] | 33.2 [1.49] | 1.000 |
| Platelet | 275 [154] | 244 [118] | 273 [146] | 318 [204] | 1.000 |
| **Malaria smear** |  |  |  |  |  |
| Peripheral blood parasite detection | 1 (1) | 1 (0.7)** | 0 (0) | 0 (0) | 1.000 |
| **Urinalysis** |  |  |  |  |  |
| Urine glucose | 17 (11) | 5 (3.4) | 7 (4.7) | 5 (3.4) | 1.000 |
| Bilirubin | 112 (75) | 29 (19.5) | 63 (42.3) | 20 (13.4) | 1.000 |
| Ketones | 75 (50) | 20 (13.4) | 39 (26.2) | 16 (10.7) | 1.000 |
| Specific gravity*** | 110 (74) | 34 (22.8) | 52 (34.9) | 24 (16.1) | 1.000 |
|  | 26 (17) | 5 (3.4) | 19 (12.8) | 2 (1.3) |  |
| Occult hematuria | 31 (21) | 8 (5.4) | 17 (11.4) | 6 (4) | 0.963 |
| Urine pH | 6.41 [1.05] | 6.59 [1.10] | 6.39 [0.97] | 6.22 [1.16] | 1.000 |
| Proteinuria | 114 (77) | 31 (20.8) | 62 (41.6) | 21 (14.1) | 1.000 |
| Urobilinogen | 56 (38) | 12 (8.1) | 31 (20.8) | 13 (8.7) | 1.000 |
| Nitrites | 30 (20) | 6 (4) | 20 (13.4) | 4 (2.7) | 1.000 |
| Leukocytes esterase | 37 (25) | 11 (7.4) | 20 (13.4) | 6 (4) | 1.000 |

† Statistics for G1 (mild progressors), G2 (intermediate progressors), and G3 (severe progressors) performed by Kruskal-Wallis test and Fisher's exact test with FDR.
\* mean [S.D.].
\*\* No. of positive (%).
\*\*\* The top row shows the number of people in specific gravity level 2 and the bottom row shows the number of people in specific gravity level 3.

### Quantifying and stratifying lesion transition dynamics of MPXV-infected patients

Generally, mpox lesions can be classified into seven distinct stages: macules, papules, vesicles, pustules, umbilication, scabbing, and desquamation [27, 28, 32]. As depicted in the distribution of lesion counts at various stages in **Extended Data Fig 2**, the average lesion count at the macule, papule, vesicle, umbilicated, and pustule stages reached zero by approximately 20 days after onset of the first lesion, whereas the count at the scab and desquamation stages persisted even beyond 40 days post-onset (**Fig 1A**). Nevertheless, there was significant individual-level variation in the number of lesions and in the temporal dynamics of lesion transition (**Extended Data Fig 2**).

**Figure 1.**
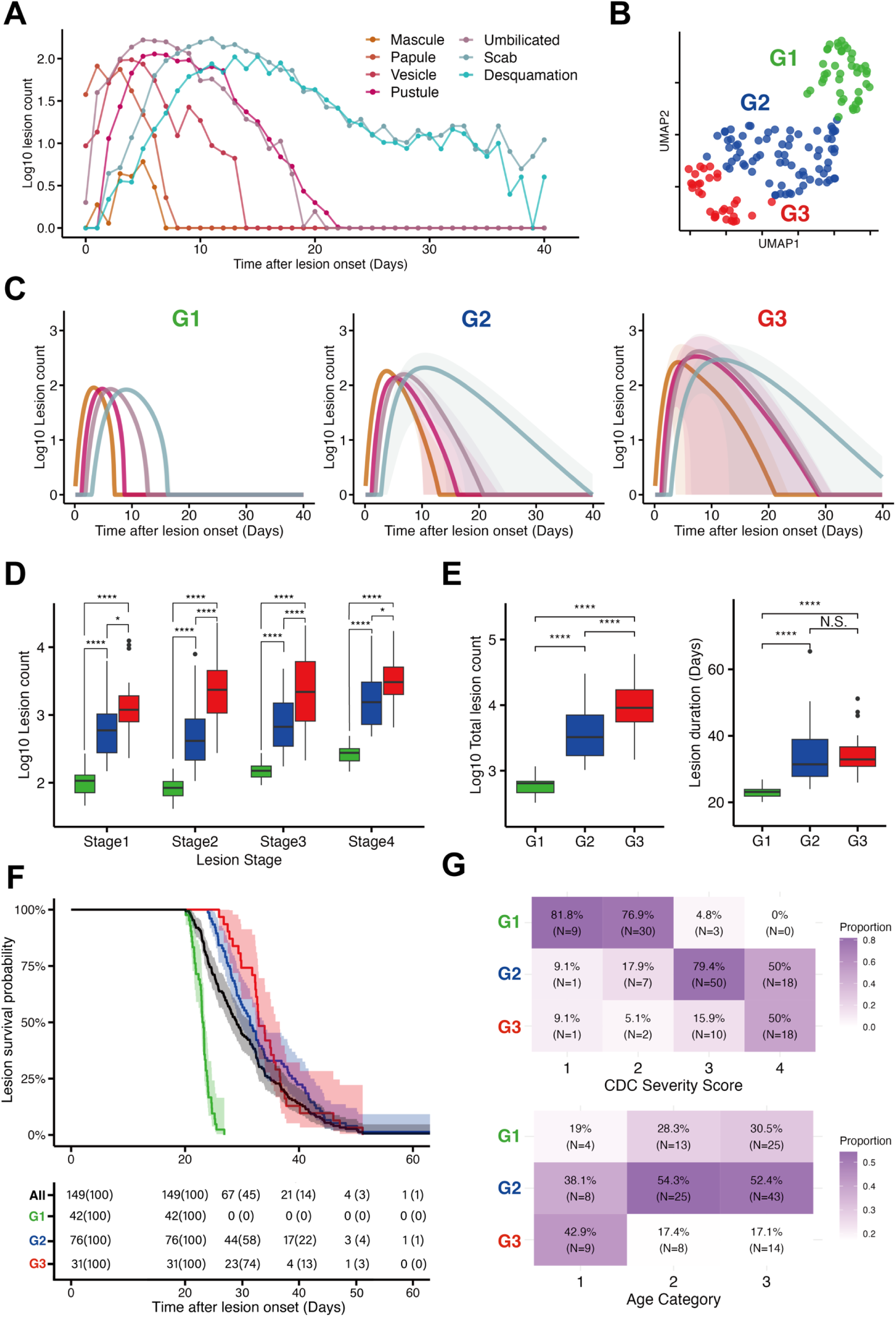
Stratification and characterization of lesion transition dynamics: **(A)** Time course plot of the average lesion count for each stage (i.e., macule, papule, vesicle, pustule, umbilicated, scab, and desquamation) across all patients. **(B)** UMAP of stratified lesion transition dynamics based on the estimated parameters. Data points represent individual participants and are colored according to group (i.e., G1, G2, and G3: green, blue, and red). **(C)** The time-course patterns highlighted by Partial Least-Squares Discriminant Analysis (PLS-DA), which discriminates each group from the others. All lesion count values for each time were reconstructed to highlight the difference between lesion stages. The mean (solid line) and mean ± 1 standard deviation (shaded area) calculated based on these reconstructed values are shown, colored according to stage (i.e., *L*_1_, *L*_2_, *L*_3,_ and *L*_4_: orange, magenta, light purple, and cyan). **(D)** Total number of lesions at each stage for each stratified group, calculated based on the reconstructed lesion transition dynamics. **(E)** Number of lesions (left) and period from lesion onset to disappearance (right), calculated based on reconstructed lesion dynamics. For **(D)** and **(E)**, statistical significance was calculated using the pairwise Mann-Whitney U test. Also, p-values were corrected by Bonferroni’s method (NS.: p-value > 0.05, ∗: p-value ≤ 0.05, ∗∗: p-value ≤ 0.01, and ∗∗∗: p-value ≤ 0.001, respectively). The statistical significance of the difference is indicated on the top of the plots. **(F)** Kaplan-Meier survival curves of lesion disappearance for each group based on reconstructed lesion dynamics. Solid lines and shaded areas indicate the survival probabilities and 95% confidence intervals, respectively. The green, blue, red, and black colors correspond to G1, G2, G3, and the total group, respectively. The table below the figure shows the number and percentage of survivors in each group at each time point. **(G)** Confusion matrices on the relationship between the stratified group and CDC severity score (top panel, individuals with <25, 25-99, 100-499, and >=500 lesions) as well as between stratified group and age category (bottom panel, individuals <5, 5-11, and >=12 years old).

To quantify the variation among MPXV-infected patients (e.g., **Extended Data Fig 2**), we used a mathematical model that describes the dynamics of lesion transition (see **Methods** in detail). For model fitting, we merged the macule, papule, and vesicle stages into one stage, owing to the limited number of lesions observed at each stage and the lack of clinical relevance in distinguishing between these three stages. Additionally, considering that the infectious period persists until all scabs (or crusts) have fallen off [28], we excluded the desquamation stage from our analysis. Consequently, we modelled lesion counts using the compartments *L*_1_, *L*_2_, *L*_3_, and *L*_4_, in which *L*_1_ represents lesions in the macule, papule, and vesicle stages, and *L*_2_, *L*_3,_ and *L*_4_ represent lesions in the pustule, umbilicated, and scab stages, respectively (**Extended Data Fig 3**). We then characterized the transition dynamics among these stages at the individual level. We reconstructed the lesion transition dynamics for each participant in our cohort over a period of approximately 40 days following the onset of lesions (see **Methods** for details). The individual-specific model fits are shown in **Fig S1** and the estimated parameters are summarized in **Table S1**.

We performed an unsupervised clustering analysis using a dissimilarity-based random forest clustering approach [37, 38] to stratify the time-course patterns of lesion transition based on estimated individual-level parameters (see **Methods** in detail). The number of clusters optimized by the algorithm was three (i.e., three groups of patients; G1, G2, and G3). **Fig 1B** represents a two-dimensional Uniform Manifold Approximation and Projection (UMAP) embedding of these three groups, which clearly shows that G1 is distinct from the other groups. Using a different color for each group, we also plotted the reconstructed individual transition dynamics in **Extended Data Fig 4**. The corresponding projected time course of each group in **Fig 1C** shows that the lesion transitions between stages progressed rapidly among individuals in G1 (N=42), whereas they were slower in the G3 participants (N=31). On the other hand, individuals in G2 (N=76) showed intermediate patterns, even though the dynamics of the scab disappearance stage (the cyan curve) were comparable to that in G3. These patterns suggest substantial variation in the infectious period of MPXV-infected patients, consistent with our recent findings of substantial individual variations in the duration of viral shedding in different specimens of MPXV clade IIb B.1 [39]. Our statistical tests showed that individuals in G1 and G3 had significantly smaller and larger numbers of lesions for all stages, respectively, compared with individuals in the other groups (**Fig 1D**). Additionally, we observed statistically significant differences in total lesion counts and lesion duration (defined as time between onset and lesion disappearance) between patient groups, except between G2 and G3 in lesion duration (**Fig 1E**). These findings demonstrate that G2 and G3 are distinguished by the total number of lesions as well as by the lesion counts in each stage, rather than by lesion duration. Hereafter we classify G1 as “mild progressors” and G3 as “severe progressors” in our analysis.

To describe the difference in timing of lesion disappearance, we calculated the probability of detectable lesions over time by using the model with estimated parameters for each group (**Fig 1F**). We found that individuals in G1 had significantly shorter periods of lesion symptoms: there was a difference in lesion disappearance between G1 vs G2, and between G1 vs G3 (p<0.0001 by log-rank test for both). In the total group (i.e., a group of all analyzed cases), the probability of detectable lesions was 55% (95% CI: 48%-64%) at 31 days after lesion onset, consistent with a reported median duration of experiencing apparent lesions among mpox cases in the 2022 outbreak [40]. All lesions in G1 patients had disappeared at 31 days after lesion onset, whereas the corresponding probabilities in G2 and G3 were 51% (95% CI: 41%-64%) and 71% (95% CI: 57%-89%), respectively. In all stratified groups, the probability of detectable lesions was less than 10% by 44 days after lesion onset.

Additionally, we compared our stratified groups with two major categorizations of MPXV-infected patients to assess reliability from a clinical standpoint: "CDC [Centers for Disease Control and Prevention] lesion severity categories” [25] based on the total number of lesions on admission day (i.e., individuals with <25, 25-99, 100-499, and >=500 lesions are categorized as 1, 2, 3, and 4, respectively) and "age categories” [27] (individuals <5, 5-11, and >=12 years old are categorized as 1, 2, and 3, respectively) (**Fig 1G**). Both categories were based on clinical investigations. We observed that CDC severity categories generally aligned with our stratified groups (e.g., patients in G1 typically had fewer lesions at admission, and those in G3 had more lesions). In our logistic regression analysis, using the number of lesions at symptom onset as the predictor and the stratified group as the outcome, we obtained high ROC-AUC values: 94% for G1 and 82% for G3. Maximizing the Youden Index from the ROC curves, we calculated cutoff values of 107 and 281 lesion counts with 62% and 61% specificity and 87% and 72% sensitivity for predicting mild and severe progressors, respectively. Notably, the cutoff value for G1 (i.e., 107 lesion counts) closely matched the boundary between CDC lesion severity categories 2 and 3 (100 lesion counts) [28]. In terms of age categories, children under 5 years old generally exhibited a longer lesion duration and a larger number of lesions, which aligns with previous reports indicating severe lesion symptoms in young children [17, 27, 41].

### Viral load at lesion onset may define severity on lesion symptoms

While the CDC lesion severity categories are useful, it is practically (and ethically) challenging in clinical settings to always count all lesions appearing on the entire body of a patient [42]. Recent studies have suggested that the diagnosis of small lesions is difficult even for clinicians and quite subjective [43]. To provide more objective support for the mpox diagnostics, we explored the relationship between the lesion transition dynamics and individual-level viral load, as our recent study suggested that viral load in lesions is positively associated with the infectious period of a patient (i.e., the duration of having culturable MPXV) for clade IIb MPXV [33, 34, 44].

In our cohort, although we measured the viral load in peripheral blood, pharyngeal swabs, and scabs [27] (see **Methods**), our focus was on the viral load in peripheral blood because the other viral loads are not routinely measured (typically once per patient in most cases). All patients (N=151; **Extended Data Fig 1**) showed a decaying viral load profile (**Fig 2A**), since the viral load is likely to have already peaked before the first measurement (i.e., before patients present to the hospital) [27]. Therefore, we used a decay model to reconstruct the best-fit viral load profile for each case, taking into account inter-patient heterogeneity (see **Methods**). In **Fig 2B**, we depict the reconstructed individual dynamics aligned with the stratified groups (i.e., G1, G2, and G3). We compared four key features derived from the viral dynamics among the groups in **Fig 2C**: viral load at lesion onset and at disappearance (when scab count falls below 1), total viral load (defined as the AUC of viral load above 10^−2^ genomes/mL), and the viral clearance rate. Interestingly, we found that the onset viral loads and total viral load in G1 were significantly lower than those in G3, whereas the viral loads at lesion disappearance in G1 were significantly higher than those in G2. Conversely, there was no apparent difference in the clearance rate among the groups. On an individual patient level, we observed positive correlations for the two main quantified lesion dynamics features (lesion duration and total lesion counts in **Fig 1E**) with the onset viral load in **Fig 2D** (0.39 and 0.49 Pearson’s correlation coefficient, respectively, with a p-value of 0.001 in both cases). These findings suggest that the onset viral load may serve as a potential biomarker for predicting lesion transition patterns and severity of illness over the course of infection (discussed below). Additional correlation analyses between lesion dynamics and viral load dynamics are illustrated in **Extended Data Fig 5**.

**Figure 2.**
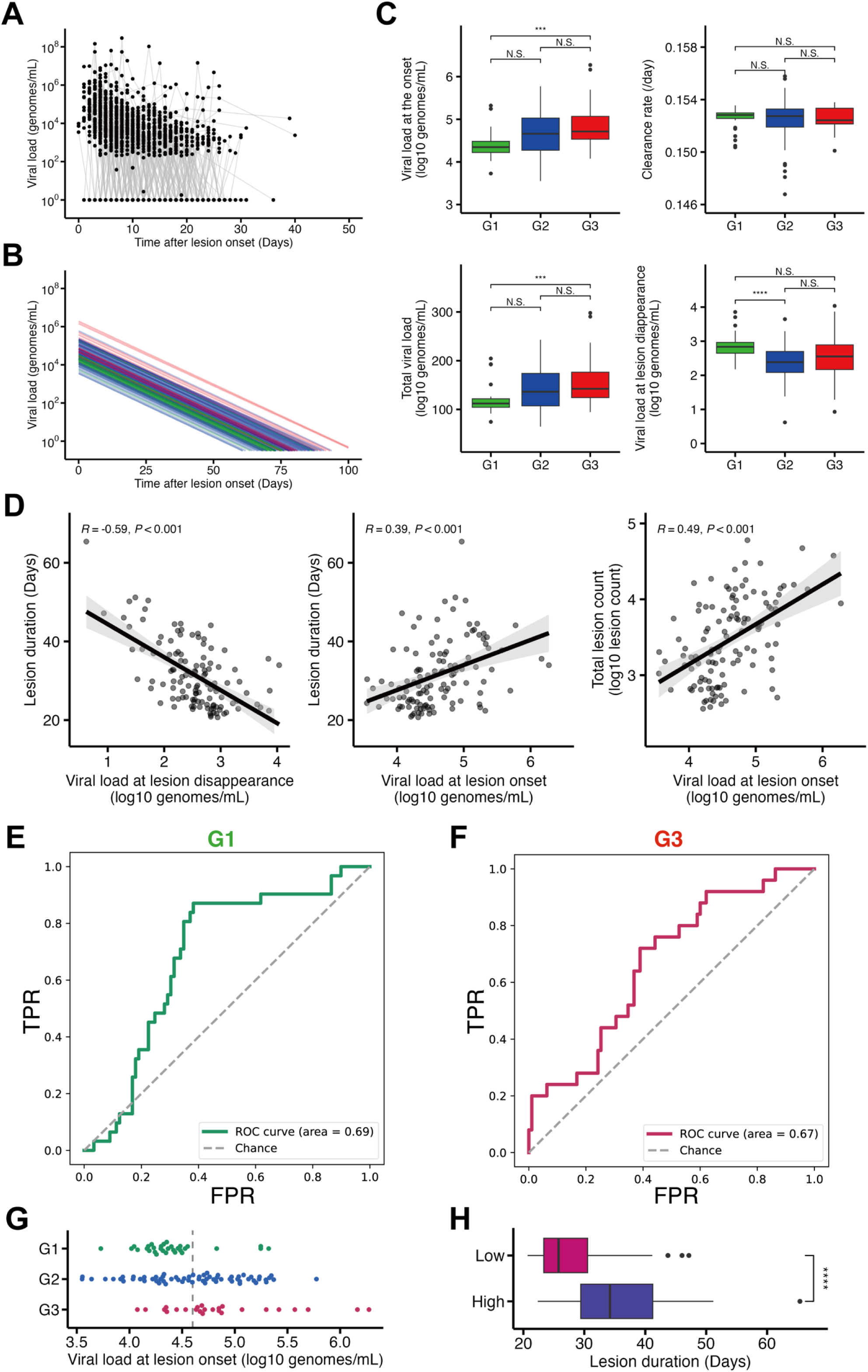
Relationship between lesion transition dynamics and peripheral blood viral load: **(A)** The measured individual decay dynamics of mpox viral load (i.e., genomic DNA) in peripheral blood samples obtained from the DRC cohort. **(B)** The individual-level mpox decay viral load, reconstructed using the mathematical model, plotted aligned with the stratified groups (i.e., G1, G2, and G3: green, blue, and red). **(C)** Comparison of viral load at onset, viral clearance rate, total viral load in peripheral blood, and viral load at lesion disappearance among the stratified groups. **(D)** Correlations between the features of individual-level lesion transition dynamics (i.e., the lesion duration and total lesion counts) and those of individual-level decay viral load dynamics (i.e., the viral load at lesion onset and disappearance). **(E)(F)** The ROC curves for a generalized linear model (GLM) logistic regression with the viral load at lesion onset for predicting stratified group G1 or not and G3 or not, respectively. The corresponding ROC-AUCs are displayed in the bottom right of each panel. FPR and TPR mean false positive rate and true positive rate, respectively. **(G)** Swarm plot of the viral load at onset among the stratified groups. The dashed line indicates the threshold value (4.6 log_10_ genomes/mL). **(H)** Comparison of lesion duration between two groups classified based on the threshold value (Low: below 4.6 log_10_ genomes/mL, High: above 4.6 log_10_ genomes/mL).

Finally, we explored whether an individual’s risk group (G1, G2, or G3) could be predicted by the onset viral load in peripheral blood. We used a logistic regression analysis with the onset viral load as the predictor variable and the stratified groups as the outcome variable. Although we could not attain a high ROC-AUC for predicting G2 (41%), which exhibits intermediate patterns, we did achieve relatively higher ROC-AUCs of 69% for G1 (defined as mild progressors) and 67% for G3 (defined as severe progressors) as shown in **Fig 2EF**. Similarly, based on the Youden Index, we calculated cutoff values for the onset viral load of 4.557 and 4.619 (log_10_ genomes/mL) with 61.8% and 61.1% specificity and 87.1% and 72.0% sensitivity for predicting mild and severe progressors, respectively. Interestingly, these two cutoff values were very close, and therefore we henceforth use a common threshold of an onset viral load of 4.6 (log_10_ genomes/mL) instead. **Fig 2G** shows that the threshold generally distinguishes between G1 and non-G1 (i.e., mild or non-mild progressors) as well as between G3 and non-G3 (i.e., severe or non-severe progressors), but not the intermediate pattern (i.e., G2). Notably, the threshold viral load significantly differentiated the duration of lesions (p<0.0001 by Mann-Whitney U-test): 28.0 and 35.6 days on average are required until lesion disappearance for patients with an onset viral load below and above 4.6 (log_10_ genomes /mL), respectively (**Fig 2H**).

### Characterizing mild and severe progressors by clinical laboratory data

In this section, we further explored potential biomarkers for use in predicting mild and severe progressors by incorporating additional clinical laboratory data. Using the clinical laboratory data obtained from our cohort [27] (see **Methods**), we annotated the 149 patients with hematology analysis, urinalysis, and clinical chemistry on the day of lesion onset (**Table 1** and **Extended Data Fig 1**). We examined the correlation between the stratified group based on estimated lesion transition patterns and the 41 variables (summarized in **Table 1**). Each factor was compared between the three groups by Kruskal-Wallis and Fisher’s exact test where the p-values were corrected by the false discovery rate (FDR). However, we found that only the white blood cell count differed significantly among the stratified groups (i.e., adjusted p-value of less than 0.05).

Next, we investigated whether the inclusion of the clinical laboratory data alongside the onset viral load improved the performance of the model predictions in **Fig 2EF**. We here used a supervised machine learning algorithm (Light Gradient Boosting Machine [LightGBM]) to predict the group from the laboratory data of 120 patients in our cohort (see details in **Methods**). Despite the inclusion of additional data, we achieved similar ROC-AUCs, specifically for G1 (70%) and G3 (64%), as depicted in (**Fig 3AB**), along with a lower ROC-AUC for G2 (53%). Interestingly, the combination of viral load data and clinical laboratory information did not significantly improve predictability for either mild or severe progressors.

**Figure 3.**
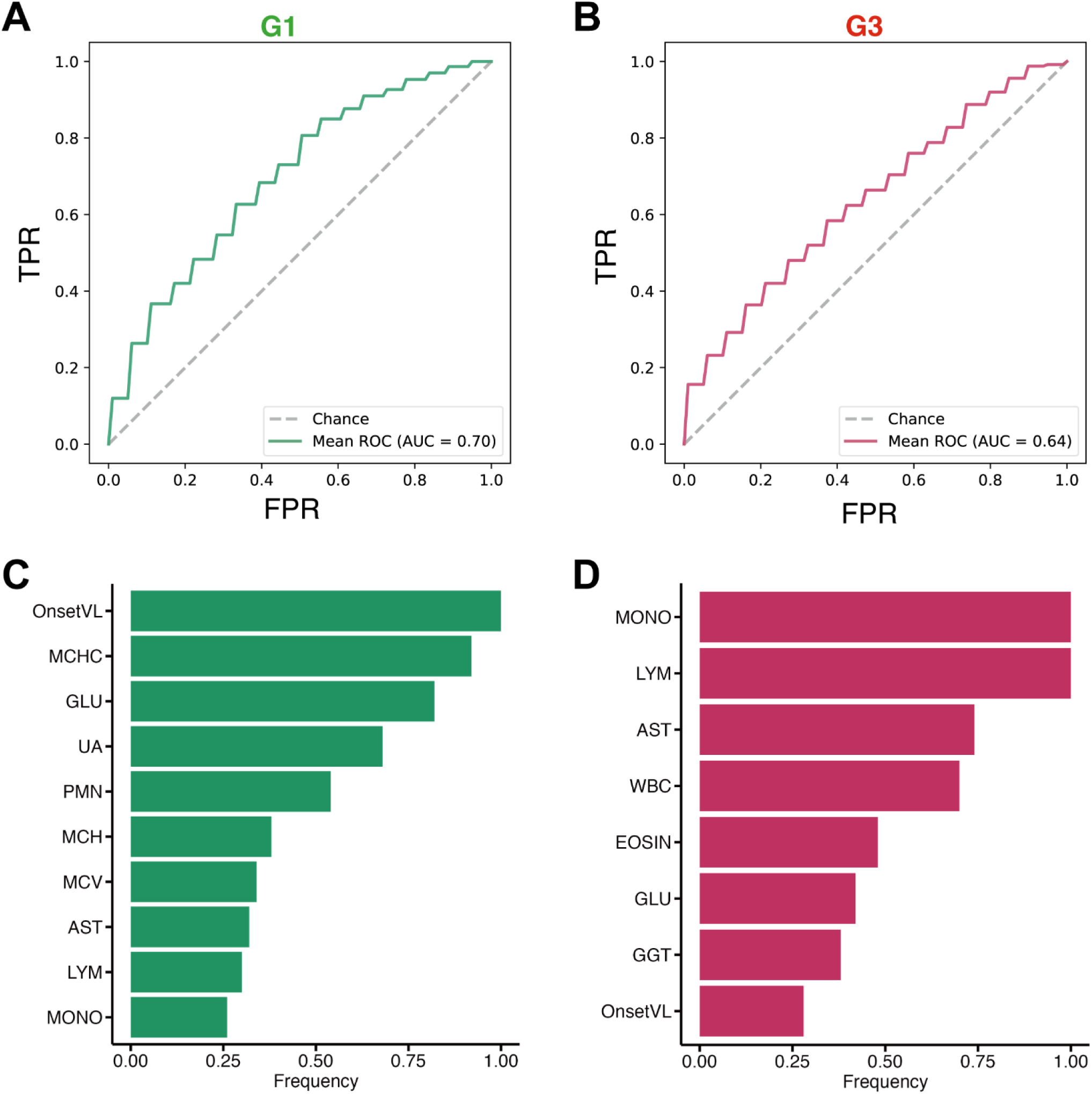
Clinical characterization of stratified groups: **(A)** and **(B)** depict the ROC curves of LightGBM classifiers trained to predict either G1 (i.e., mild progressor) or not, and G3 (i.e., severe progressor) or not, respectively. The corresponding ROC-AUCs are calculated and displayed in the bottom right of each panel. **(C)** and **(D)** show bar plots of how often a feature was selected among seeds as an indicator of selection stability. Selected features are shown with a frequency greater than 0.25. OnsetVL: viral load at lesion onset, MCHC: mean corpuscular hemoglobin concentration, GLU: blood glucose level, UA: uric acid, PMN: neutrophils, MCH: mean corpuscular hemoglobin, MCV: mean corpuscular volume, AST: aspartate aminotransferase, LYM: lymphocytes, MONO: monocytes, WBC: white blood cell, EOSIN: eosinophils, GGT: gamma glutamyl transferase.

Finally, concentrating on G1 (mild progressors) and G3 (severe progressors), we examined features (other than the onset viral load) that predict which of these two non-intermediate groups’ individuals belong to. In **Fig 3AB**, we selected features for our model to enhance classification accuracy and efficiency by eliminating redundancy. Specifically, the feature selection was based on the importance of SHapley Additive exPlanations (SHAP) values [45], which were added to the LightGBM model in order of significance (see **Methods**). The features with the highest ROC-AUC in the training dataset were then chosen. Following feature selection, the frequencies of the features selected over 50 iterations [46] for G1 and G3 are shown in **Fig 3CD**. Additionally, the distribution of corresponding features between G1 as opposed to non-G1, and G3 as opposed to non-G3, is depicted in **Extended Data Fig 6**, respectively. The frequency lists of selected features for both G1 and G3 included the onset viral load as the 1st and 8th feature, respectively. This again supports the importance of onset viral load in predicting the severity of lesion symptoms. Regarding other biomarkers, we found that low viral load and low AST (which indicates absence of liver damage at onset) were weakly associated with mild progressors. In contrast, for the severe progressors, the most relevant biomarkers were high WBC and AST, which implies elevated inflammatory response and liver dysfunction at onset. These clinical findings suggest that inflammatory response and virus-induced organ damage at onset may be linked to disease progression.

## Discussion

In the present study, using a large cohort of mpox cases in the DRC from our previous study [27], we have further investigated the dynamics of lesion symptoms at the individual patient level. We have identified groups of mild and severe progressors (G1 and G3); the G1 group was characterized by a lower lesion count and more rapid disappearance of lesions, whereas the G3 group exhibited a higher lesion count and slower disappearance of lesions. Our stratification also identified age as a key demographic characteristic affecting lesion dynamics, in accordance with previous studies which suggested that younger age is associated with more likely severe outcomes, which determine the necessity and benefits of hospital-based care [17, 27, 41].

Limited research has been done on potential biomarkers for the severity of mpox, which could be used to monitor clinical progress and tailor treatment plans in individual patients [47]. In a recent study [48], some peptides were identified to be correlated with severity, although the plasma proteome analysis was confined to a small number of mpox patients. In contrast, our analysis with data from over 150 mpox patients suggested that the viral load in peripheral blood at lesion onset may serve as a biomarker capable of predicting severity in the early stages of lesion symptom progression. Specifically, it may be possible to differentiate between mild and severe progressors by comparing their viral load at lesion onset with a threshold value of 4.6 (log_10_ copy/mL) and also to predict the possible timing of lesion disappearance. To swiftly prioritize patients for either home treatment or hospitalization, especially for individuals who go on to experience complications [36], the onset viral load has practical advantage, as viral load data can be obtained during routine practice in clinical settings, such as when confirming a case with PCR testing. In the future, this research may lead to remote diagnosis of Mpox infections in areas with difficult access to medical care, where prognosis and treatment methods can be determined based solely on images of skin lesions.

Furthermore, adaptive treatments that account for the stratified time-course patterns of lesion transition become feasible if patients can be classified into severity groups shortly after being diagnosed with MPXV infection (i.e., the date of symptom onset). For patients classified in group G1 (i.e., mild progressors), tecovirimat may represent an appropriate treatment, whereas for those in G3 (i.e., severe progressors), combination treatment using tecovirimat with cidofovir, brincidofovir, or vaccinia immune globulin intravenous (VIGIV) may be reasonable [49, 50]. Establishing an early treatment plan is crucial for improved patient outcomes and efficient health care delivery.

Our study has several limitations. First, our analysis relied on patient data from the DRC cohort of cases infected with clade I MPXV, and thus caution should be taken when using our findings to inform treatment or interventions during outbreaks of other clades or orthopoxviruses, such as the dominant clade MPXV in 2022 (i.e., clade IIb). Delays between infection and recognizable symptom onset, or between symptom onset and confirmation/hospitalization may differ by clade/virus [51] (and potentially by mode of transmission [52, 53]), and timeliness is crucial when measuring onset viral load data. Overall, our estimates of disease progression after lesion onset were mostly consistent with epidemiological findings observed in 2022 [3, 26, 34], and the added value of our analysis was to characterize the individual-level disease progression and quantify the duration of symptoms by the level of severity. Our results also provide timely evidence for the control of the clade I mpox outbreaks currently surging in the DRC and surrounding countries [54], which have a higher case fatality ratio than outbreaks due to clade IIb [6, 7, 17, 18]. Second, further investigation is needed on the association between lesion counts at each stage and the mpox viral load in peripheral blood, both in patients who have not been treated with antiviral drugs and in those receiving medications such as tecovirimat, cidofovir, and brincidofovir [49, 55]. This is particularly relevant in outbreak settings where these drugs may be administered to infected individuals and used prophylactically. For example, although tecovirimat is currently available only under emergency authorization, clinical trials assessing its efficacy in the treatment of mpox are underway [56]. In fact, case studies of patients treated with tecovirimat show anecdotal improvement in symptoms and viral clearance [57, 58]. Finally, in our machine learning analysis, clinical laboratory data were available for only 120 patients because of missing measurements, and the data needed to be further divided into 80% training and 20% test datasets. The limited sample size may explain why we did not achieve high ROC-AUCs despite the inclusion of the clinical data. These challenges highlight the need for larger datasets to be collected.

In conclusion, this study presents empirical evidence of heterogeneity in lesion transition dynamics and symptom severity among mpox cases. It also suggests that the viral load in peripheral blood at lesion onset may be linked to this heterogeneity. Given the absence of currently available sensitive, simple and rapid biomarkers, the onset viral load could potentially be used as a predictor of mpox skin lesion severity. Identifying risk factors for severe symptoms at the individual level is crucial for improving treatment strategies and public health interventions against current and future outbreaks of mpox.

## Methods

### Ethics statement

The prospective observational study of the clinical natural history of human MPXV infections at the remote Kole Hospital in the rainforest of the Congo River basin of the DRC, which was conducted from March 2007 until August 2011, was reviewed and approved by the Human Use Committee of the United States Army Medical Research Institute of Infectious Diseases (FY05-13) and the Headquarters, United States Army Medical Research and Development Command Institutional Review Board (IRB), Frederick, MD, USA, as well as the Ethics Committee at the University of Kinshasa School of Public Health (KSPH), Kinshasa, DRC (see [27] for details). The present study was approved by the ethics committee of Nagoya University (approval number: Hc 22-07).

### Study data

The count of lesions on nine distinct skin regions and the oropharynx of each patient’s body (a total of 10 areas) was documented and reported in [27]. In this study, we used the total number of lesions for each stage (macules, papules, vesicles, pustules, umbilication, scabbing, and desquamation) by aggregating the lesion count from all areas for each day of the study. Additionally, we incorporated viral load data, measuring the amount of MPXV genomic DNA in peripheral blood, pharyngeal swabs, and scabs. Clinical laboratory data, obtained through hematology analysis, urinalysis, and clinical chemistry from the same group of patients, was also considered. Detailed information on data preparation can be found in [27].

### Modeling lesion transition dynamics

In our recent report [27], we categorized lesions into seven different stages: macule, papule, vesicle, pustule, umbilicated, scab, and desquamation. Because of the limited number of macule, papule, and vesicle lesions for the purpose of parameter estimation and the lack of clinical relevance in distinguishing between these three stages, we merged these into one stage. The notation *L*_1_(*t*) therefore represents the total number of macule, papule, and vesicle lesions at time *t* since lesion onset (i.e., *t* = 0 represents the day at which a patient reported its first lesion appearance). Additionally, considering that the infectious period persists until all scabs or crusts have fallen off [28], we considered lesion counts in the umbilicated, pustule, and scab stages as *L*_2_(*t*), *L*_3_(*t*), and *L*_4_(*t*), respectively, while excluding lesion data from the desquamation stage. Subsequently, we developed the following mathematical model to describe the dynamics of lesion transitions in MPXV-infected patients:

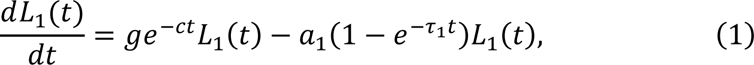

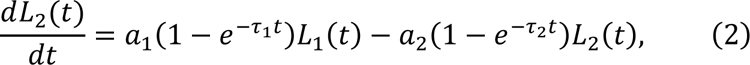

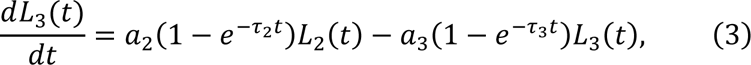

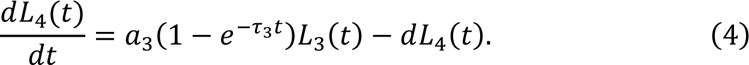

Here, we consider that the number of macule, papule, and vesicle lesions (i.e., *L*_1_) increases at per lesion rate *ge*^−*ct*^ as the infection progresses, where *g* corresponds to the maximum growth rate of the number of lesions and *c* determines the rate at which this growth rate decreases over time due to viral clearance. It is assumed that the lesion counts in *L*_1_ start to exponentially increase after *t*_0_ days post first lesion onset and *L*_1_ remains at 1 until that time (i.e., *L*_1_(*t*) = 1 and *L*_2_(*t*) = *L*_3_(*t*) = *L*_4_(*t*) = 0 for 0 < *t* ≤ *t*_0_). The differential equations above therefore apply when the time since lesion onset *t* > *t*_0_. In those equations, the number of scab lesions (i.e., *L*_4_) decreases at per lesion rate *d*, and the transition rates between the stages are denoted by *a*_*n*_(1 − *e*^−*τ_n_t*^) for *n* = 1,2,3 where *a*_*n*_ is the maximum transition rate and 1 − *e*^−*τ_n_t*^ sets the time dependence of delays between the stage transitions. We estimated the above parameters, i.e., *g*, *c*, *a*_*n*_, *d*, *t*_0_, *τ*_*n*_ for *n* = 1,2,3, by model fitting (see below).

### Modeling decay dynamics of viral load

To describe MPXV dynamics in peripheral blood, we used a simple decay model, which was previously used in a study of mpox [39]:

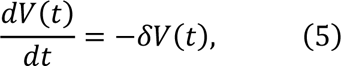

where the variable *V*(*t*) is the viral RNA load (copies/mL) at time *t* and parameter *δ* represents the viral clearance rate. Note that the variable *t* again represents time after lesion onset; *t* = 0 is thus the date on which the first lesion was identified.

### Parameter estimation

We estimated the parameters of both the lesion transition and viral load models using a nonlinear mixed-effect modeling approach, which incorporates fixed effects as well as random effects describing the inter-patient variability in parameter values. Each parameter of patient *k*, *θ*_*k*_(= *θ* × *e*^*πk*^) is decomposed as product of *θ* (the fixed effect) and *e*^*πk*^ (the random effect), where *π*_*k*_ is assumed to be drawn from a normal distribution: *N*(0, Ω). The fixed effect parameters and the standard deviations of random effects, Ω, were estimated using the stochastic approximation expectation/maximization (SAEM) algorithm, and individual parameters for each patient were then estimated using the empirical Bayes method. A right-truncated normal distribution was used in the likelihood function to account for the left censoring of the lesion count data or viral load data (i.e., when the lesion count or viral load is not detectable) [59]. MONOLIX 2021R2 (www.lixoft.com), a program for maximum likelihood estimation for nonlinear mixed-effect models, was used to fit the model to these data. We changed the initial values multiple times to avoid obtaining a local minimum of the negative log-likelihood function and to confirm the robustness of our parameter estimates.

### Unsupervised clustering and stratification of lesion transition dynamics

We extracted three key “features” of lesion transition dynamics for each individual: i) the average macule, papule, and vesicle lesion count increase rate 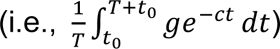 over 40 days (i.e., *T* = 40); ii) the average stage transition rates 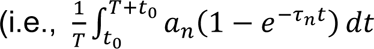 for *n* = 1,2,3), again with *T* = 40; and iii) the scab lesion decrease rate, *d*. For the purpose of stratification of the lesion transition dynamics, unsupervised random forest clustering based on each of five features was used (rfUtilities in R). The use of random forest allowed us to avoid overfitting by bootstrap aggregating (bagging) and to achieve better generalization performance [60]. Specifically, after a random forest dissimilarity (i.e., the distance matrix between all pairs of samples) was obtained, it was visualized by using UMAP in a two-dimensional plane and was stratified with spectral clustering (Python scikit-learn). The optimal number of clusters was determined using the eigengap heuristic method [61].

### Classifying the stratified groups with a logistic regression

We built a predictive model for the stratified groups using a logistic regression. In this model, we used the viral load at lesion onset as the predictor variable, with the classification of the stratified group as the outcome variable. To ensure that the model was both robust and generalizable, we subjected it to rigorous validation through a 6-fold cross-validation process. We assessed the model performance using AUC scores and reported the mean and standard deviation of these scores obtained from 50 replications [46].

### Classifying the stratified groups with a LightGBM model

Since some features showed high correlation or had missing values, we used the gradient boosting machine learning algorithm, specifically LightGBM, as our primary method of analysis [62]. LightGBM was chosen for its decision tree-based algorithm, which inherently handles the multicollinearity of predictors effectively. Given the limited number of data samples, we did not fine-tune the hyperparameters of LightGBM and the model was constructed using its default settings.

#### Data preparation

To handle missing values, we used KNNImputer [63], employing a k-nearest neighbor approach to impute missing values based on the mean of their nearest neighbors. During this process, missing values were addressed across four neighborhoods.

#### Data splitting and training algorithm

The stratification groups consisted of 42, 76, and 31 patients for G1, G2, and G3, respectively. We approached the problem as a binary classification task, where the model aimed to predict whether a given individual belonged to G1 or not (similarly for G2 and G3). The model then assigned each patient to their most likely group. We split the stratified groups into two datasets: 80% as a training dataset and 20% as a test dataset. The former was used for model training and internal validation, and the latter served as a hold-out dataset for external validation. To address class imbalance, we generated synthetic instances of the minority class in the training data using the Synthetic Minority Over-sampling Technique (SMOTE) [64].

#### Feature selection and model evaluation

Because of the propensity of machine learning models to overfit when the data contain a high number of features, feature selection was necessary. Feature selection hinged on the significance of SHAP values; we incrementally added features to the machine learning model according to their importance and selected those that yielded the highest ROC-AUC in the training dataset. The process was repeated 50 times with different pseudorandom number generator initiation seeds to account for the influence of differences in splits. Our model performance was evaluated using AUC scores, and we reported the mean and standard deviation of these scores obtained from 50 replications.

### Statistical analysis

When necessary, variables were compared among different groups using analysis of variance (the Kruskal-Wallis test and Fisher’s exact test, for more than two numerical or categorial variables, respectively) with false discovery rate, or the Mann-Whitney U test (for two numerical variables) with Bonferroni correction. All statistical analyses were performed using R (version 4.2.0).

## Data Availability

All data produced in the present study are available upon reasonable request to the authors.

## LIST OF SUPPLEMENTARY MATERIALS

**Supplementary Figure 1** | Reconstructed lesion transition dynamics by stratified group for 149 individuals

**Supplementary Table 1** | The estimated fixed and random effects for lesion transition dynamics

## ACKNOWLEDGMENTS

This study was supported in part by Grant-in-Aid for JSPS Fellows 23KJ1081 (to T.N.); Scientific Research (KAKENHI) B 23H03497 (to S.I.); Grant-in-Aid for Transformative Research Areas 22H05215 (to S.I.); Grant-in-Aid for Challenging Research (Exploratory) 22K19829 (to S.I.); AMED CREST 19gm1310002 (to S.I.); AMED Research Program on Emerging and Re-emerging Infectious Diseases 22fk0108509 (to S.I.), 23fk0108684 (to S.I.), 23fk0108685 (to S.I.); AMED Research Program on HIV/AIDS 22fk0410052 (to S.I.); AMED Program for Basic and Clinical Research on Hepatitis 22fk0210094 (to S.I.); AMED Program on the Innovative Development and the Application of New Drugs for Hepatitis B 22fk0310504h0501 (to S.I.); AMED Strategic Research Program for Brain Sciences 22wm0425011s0302; AMED JP22dm0307009 (to K.A.); JST MIRAI JPMJMI22G1 (to S.I.); Moonshot R&D JPMJMS2021 (to K.A. and S.I.) and JPMJMS2025 (to S.I.); Institute of AI and Beyond at the University of Tokyo (to K.A.); Shin-Nihon of Advanced Medical Research (to S.I.); SECOM Science and Technology Foundation (to S.I.); The Japan Prize Foundation (to S.I.). The collaboration between R.N.T. and S.I. was supported by a Royal Society International Exchange award (grant number IES-R3-193037).

## AUTHOR CONTRIBUTIONS

SI and FM designed the research. TN carried out the computational analysis. SI, FM, JWH, and PRP supervised the project. All authors discussed the research and contributed to writing the manuscript.

## INSTITUTIONAL REVIEW BOARD STATEMENT

This study was approved by the ethics committees of Nagoya University (hc22-05).

---

**Extended Data Figure 1.**
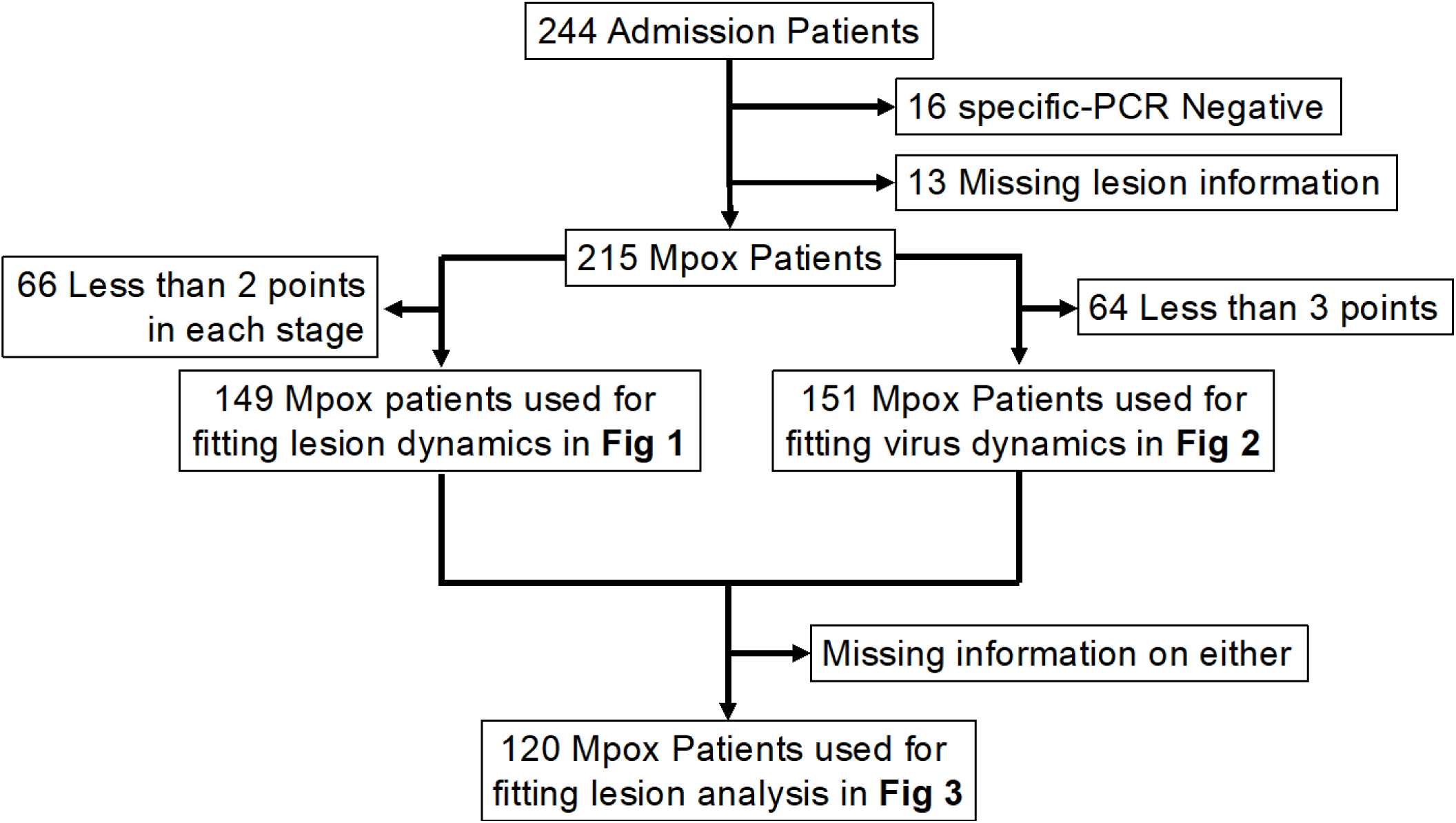
Flowchart of the cohort, along with the number of participants and inclusion criteria for our analysis.

**Extended Data Figure 2.**
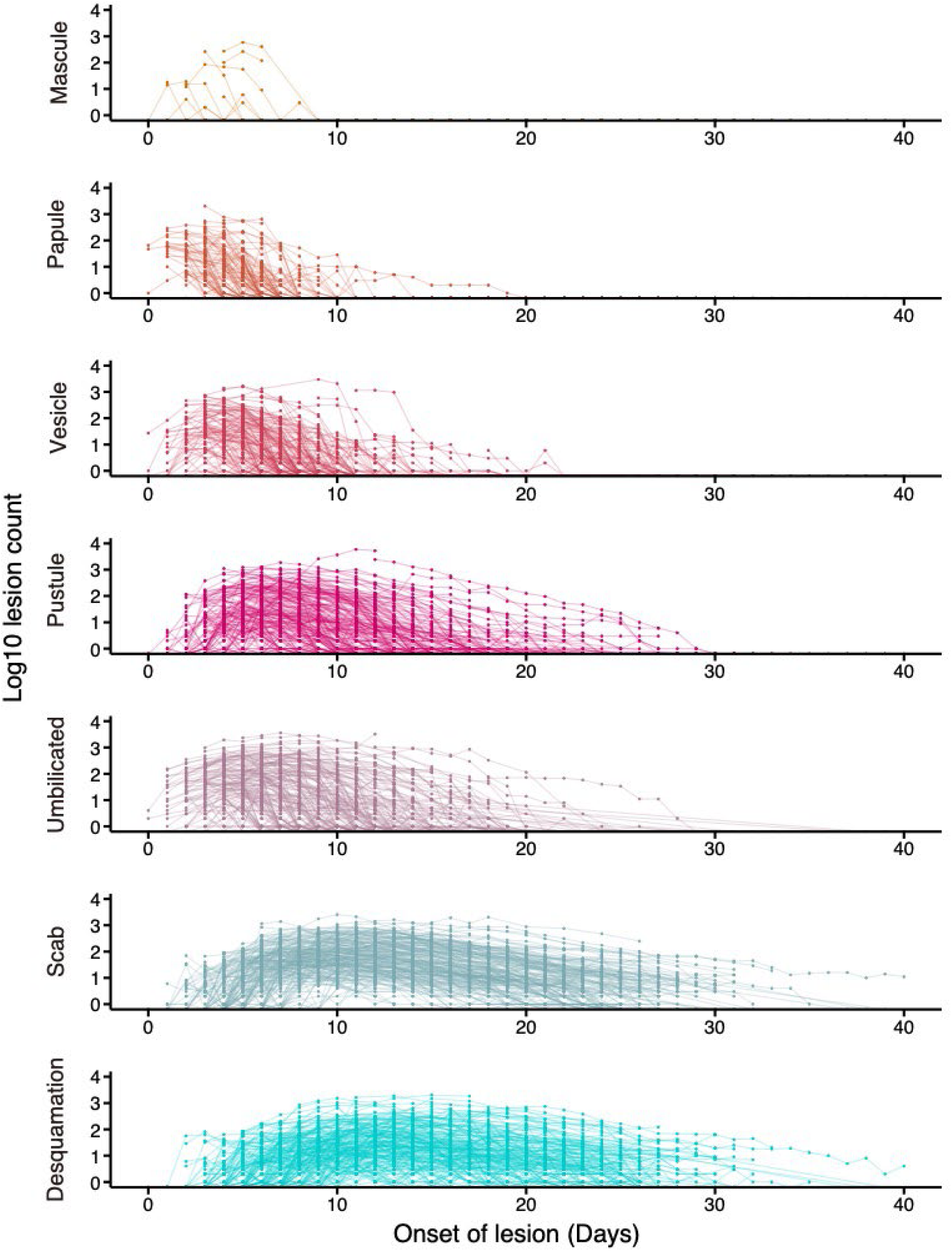
Time course distribution of the average lesion count of each stage: Time course plots of individual lesion counts for each stage (i.e., macule, papule, vesicle, pustule, umbilicated, scab, and desquamation) across all patients.

**Extended Data Figure 3.**
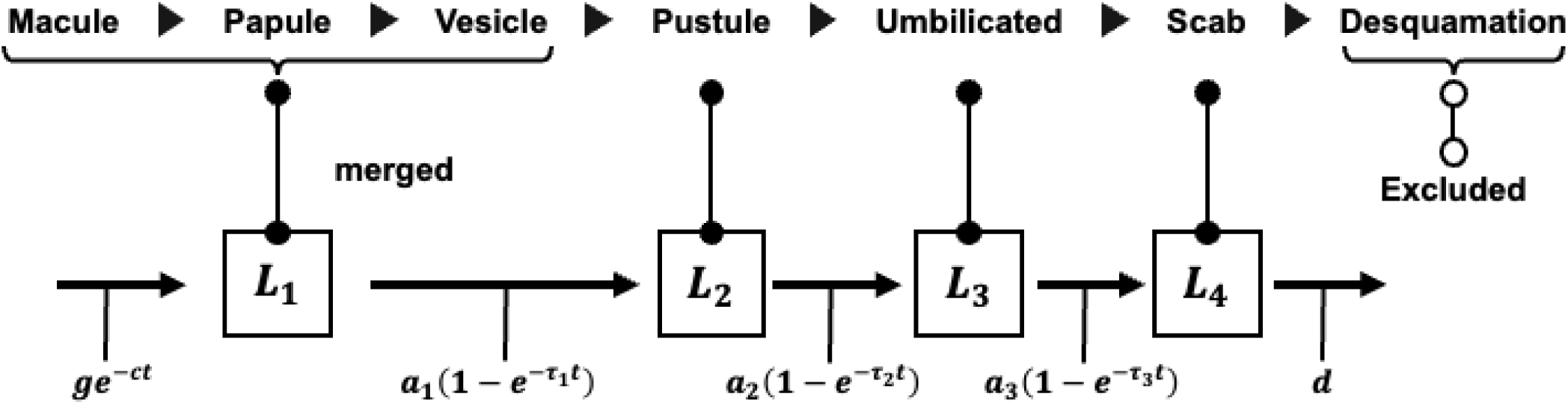
Schematic diagram of compartment model for lesion transition: The lesion counts in the macule, papule, and vesicle (i.e., these three are in one compartment, as described in the main text); pustule; umbilicated; and scab stages are defined as *L*_1_, *L*_2_, *L*_3_, and *L*_4_, respectively. It is assumed that *L*_1_ increases after *t*_0_ days post lesion onset according to a per-lesion rate *ge*^−*ct*^ as the infection progresses (with *L*_1_(*t*) = 1 for *t* ≤ *t*_0_), where *g* corresponds to the maximum growth rate of the number of lesions and *c* determines the rate at which this growth rate decreases over time. In contrast, *L*_4_ decreases according to a rate *d*, and the transition rates between the stages are denoted as *a*_*n*_(1 − *e*^−*τ_n_t*^) for *n* = 1,2,3 where *a*_*n*_ is the maximum transition rate and 1 − *e*^−*τ_n_t*^ sets the time dependence of the delay between the stage transitions.

**Extended Data Figure 4.**
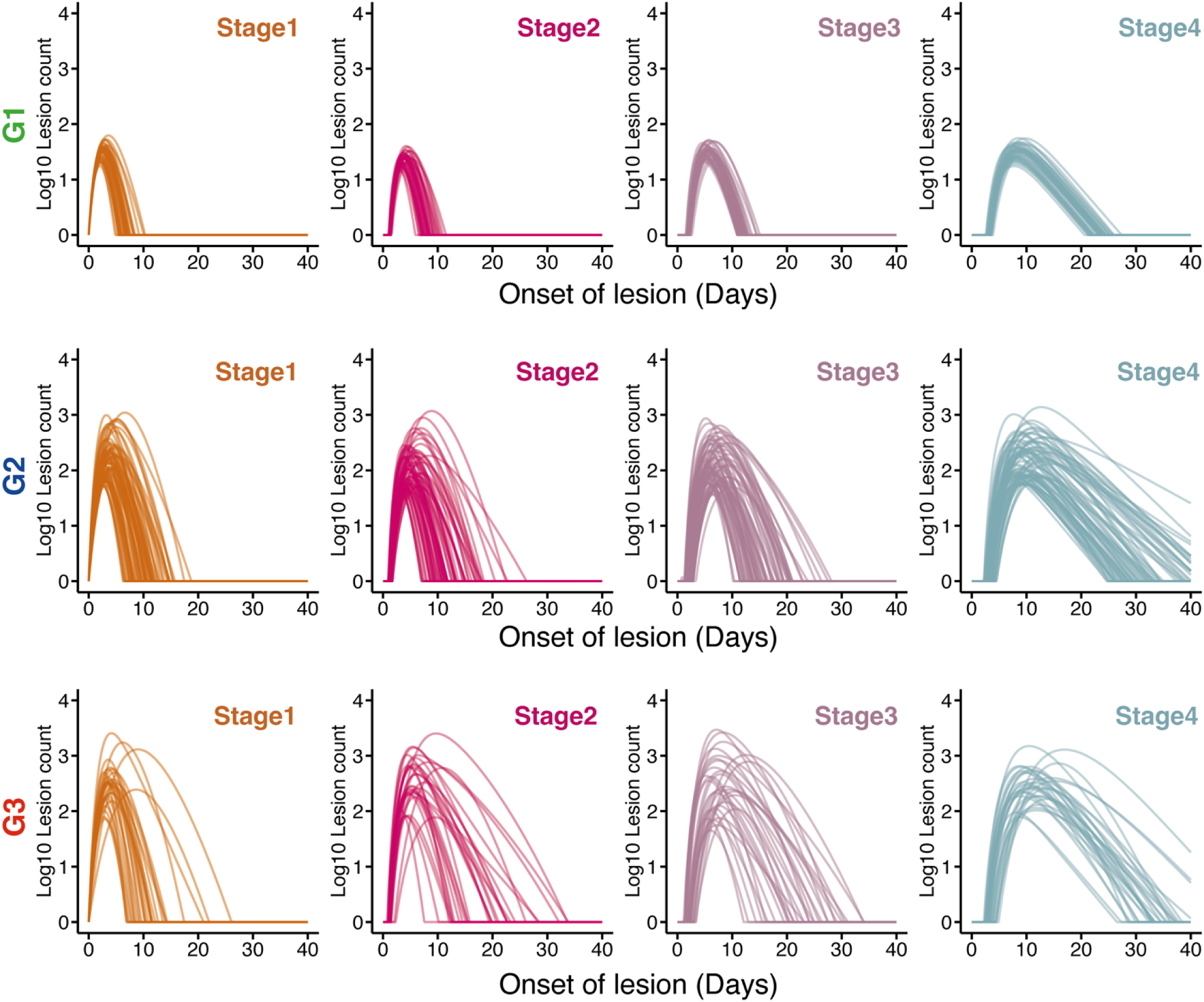
The time-course patterns of reconstructed lesion dynamics for each stage: The reconstructed individual lesion dynamics for each stage, shown for each stratified group (i.e., G1, G2, and G3).

**Extended Data Figure 5.**
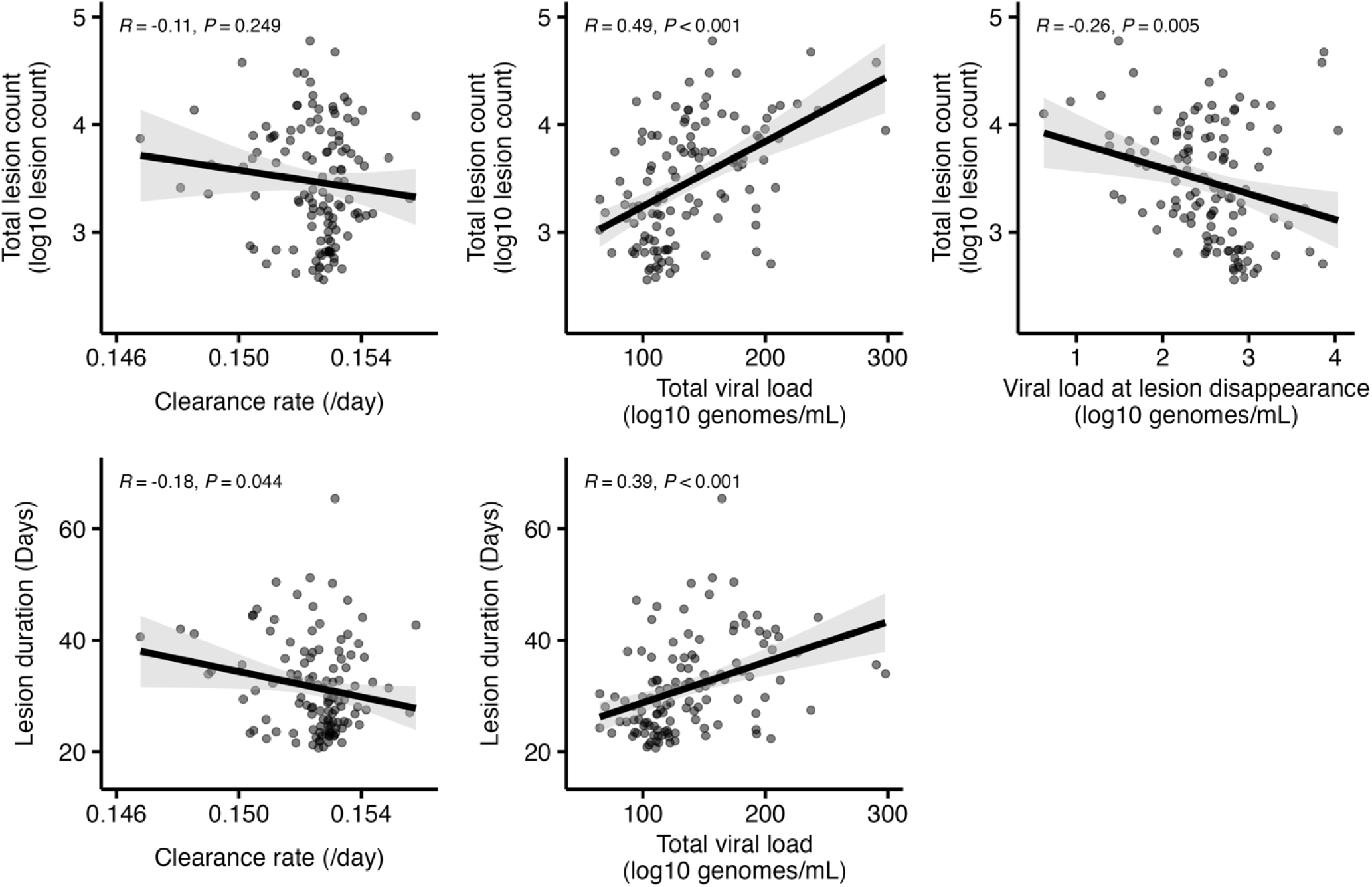
Relationship between lesion dynamics and virus dynamics: Correlations between the features of individual-level lesion transition dynamics (i.e., the lesion duration and total lesion counts) and those of individual-level decay viral load dynamics (i.e., the viral clearance rate, the total viral load and the viral load at lesion disappearance).

**Extended Data Figure 6.**
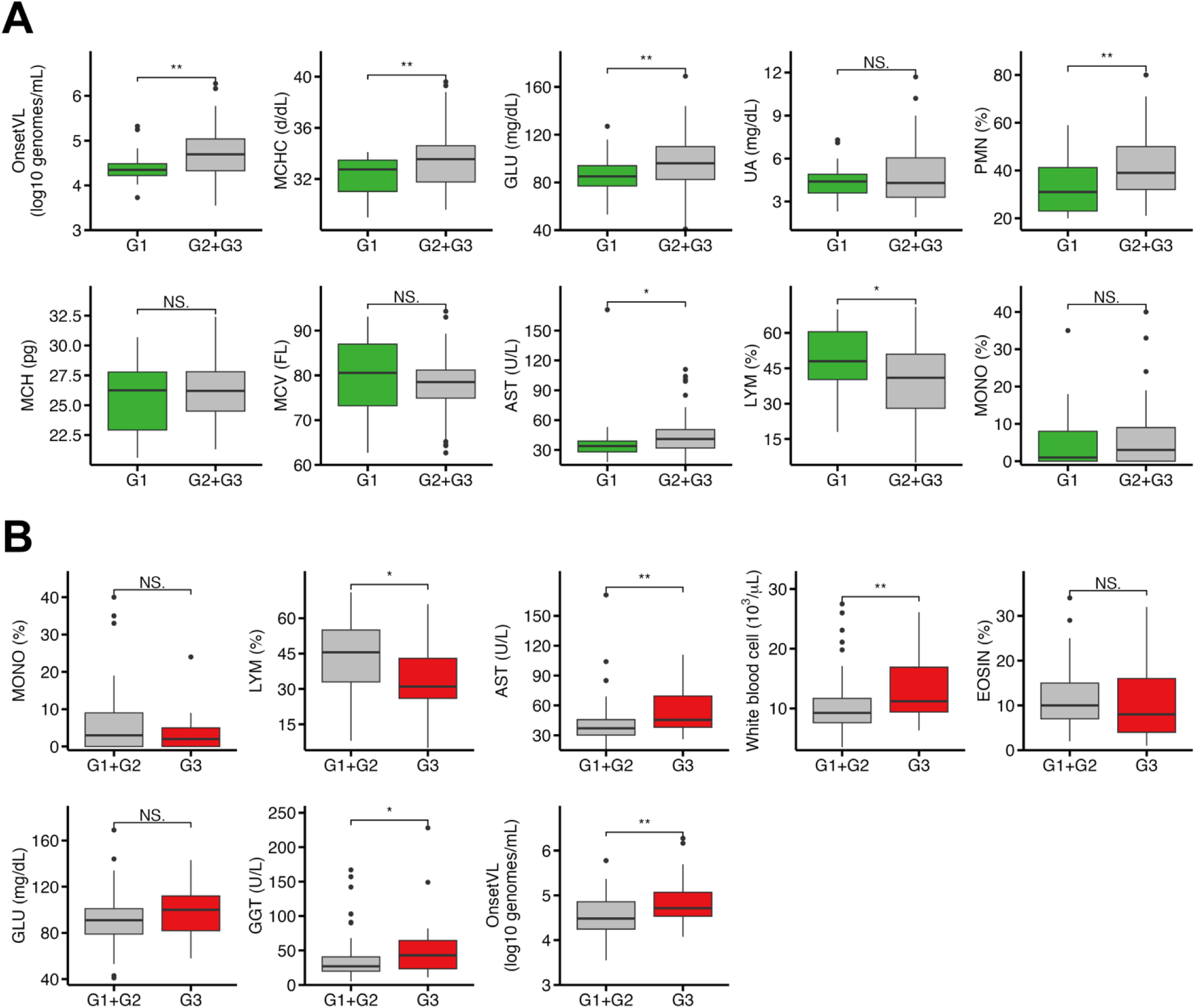
Distribution of features selected by machine learning: **(A)** and **(B)** show distributions of selected features by LightGBM with G1 or not, and G3 or not, respectively. Statistical significance is calculated using the pairwise Mann-Whitney U test. (NS.: p-value > 0.05, ∗: p-value ≤ 0.05, ∗∗: p-value ≤ 0.01, and ∗∗∗: p-value ≤ 0.001, respectively). The statistical significance of the difference is indicated on the top of the plots.

## Supplementary Information

**Supslementary Figure 1.**
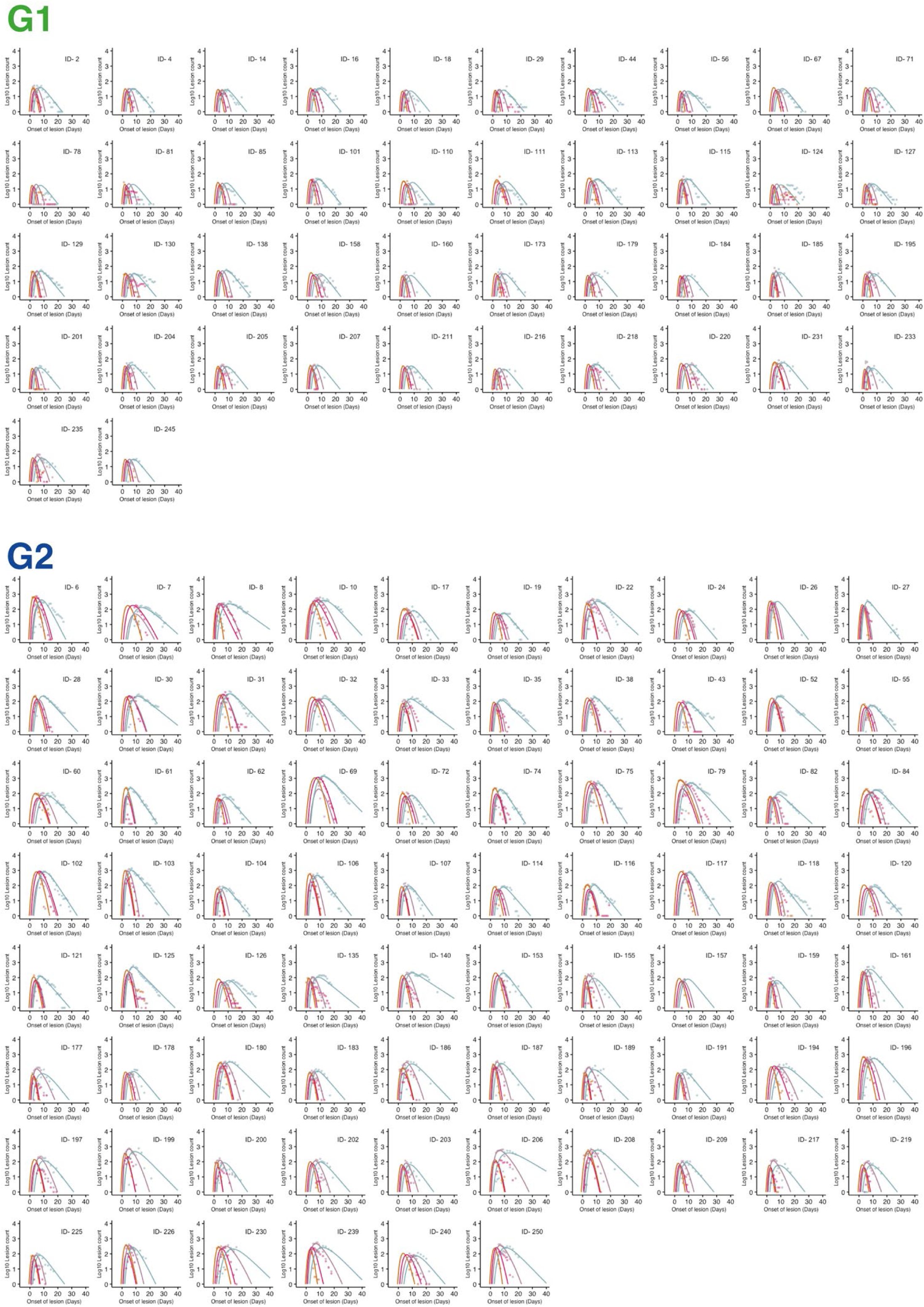
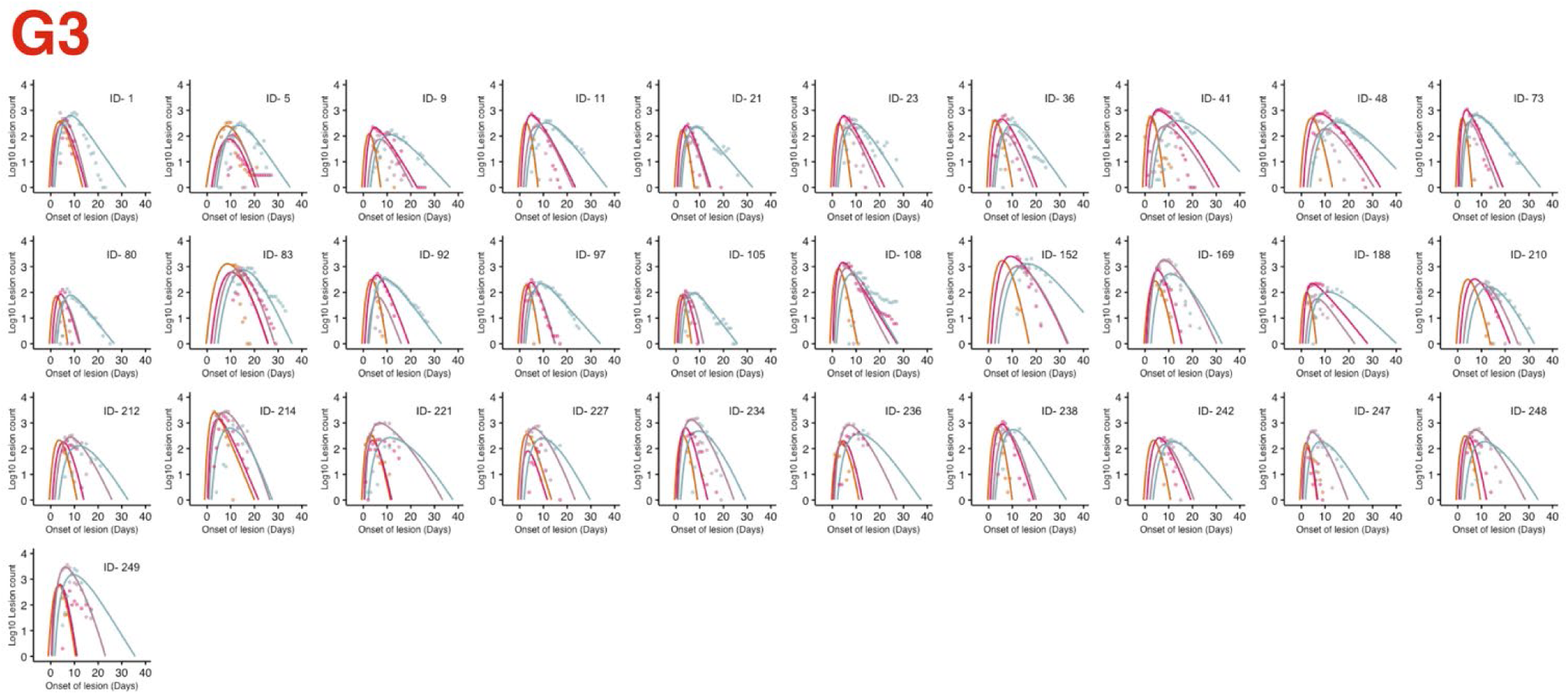
Reconstructed lesion transition dynamics for stratified groups of 149 individuals: Individual-level model fits of lesion count data using the model described in Eqs. (1-4) in the main text. The closed dots and solid curves indicate measured data and estimated lesion dynamics, respectively. Dots and curves are colored according to stages (i.e., *L*_1_, *L*_2_, *L*_3_, and *L*_4_: orange, magenta, light purple, and cyan).

**Supplementary Table 1.** Estimated fixed and random effects for lesion transition dynamics.

| Parameters | Description | Unit | $\vartheta$: Fixed effect (SE)* | $\Omega$: SD of random effect (SE)* |
| --- | --- | --- | --- | --- |
| $g$ | Maximum growth rate of the lesion | 1/day | 2.58 (0.07) | 0.18 (0.02) |
| $c$ | Decreasing rate of maximum growth rate of the lesion | 1/day | 0.53 (0.03) | 0.48 (0.04) |
| $a_1$ | Maximum transition rate from $L_1$ to $L_2$ | 1/day | 1.80 (0.11) | 0.48 (0.05) |
| $a_2$ | Maximum transition rate from $L_2$ to $L_3$ | 1/day | 1.63 (0.17) | 0.82 (0.07) |
| $a_3$ | Maximum transition rate from $L_3$ to $L_4$ | 1/day | 1.97 (0.17) | 0.68 (0.06) |
| $d$ | Decreasing rate of $L_4$ | 1/day | 0.26 (0.01) | 0.38 (0.04) |
| $t_0$ | Day at increasing lesion count | Days | 0.53 (0.03) | 0.31 (0.03) |
| $\tau_1$ | Scaling coefficient for transition delay from $L_1$ to $L_2$ | 1/day | $9.9 \times 10^{-3}$ ($0.22 \times 10^{-3}$) | 0.05 (0.02) |
| $\tau_2$ | Scaling coefficient for transition delay from $L_2$ to $L_3$ | 1/day | $9.3 \times 10^{-2}$ ($1.83 \times 10^{-2}$) | 1.56 (0.16) |
| $\tau_3$ | Scaling coefficient for transition delay from $L_3$ to $L_4$ | 1/day | $4.7 \times 10^{-2}$ ($0.38 \times 10^{-2}$) | 0.40 (0.05) |

\* The parameter for patient $k$ , $\vartheta_i (= \vartheta \times e^{\pi_k})$ is represented as a product of $\vartheta$ (a fixed effect) and $e^{\pi_k}$ (a random effect). $\pi_k$ follows the normal distribution with mean 0 and standard deviation $\Omega$ . SE: standard error.

